# Monitoring Body Composition Change for Intervention Studies with Advancing 3D Optical Imaging Technology in Comparison to Dual-Energy X-Ray Absorptiometry

**DOI:** 10.1101/2022.11.14.22281814

**Authors:** Michael C. Wong, Jonathan P. Bennett, Lambert T. Leong, Isaac Y. Tian, Yong E. Liu, Nisa N. Kelly, Cassidy McCarthy, Julia MW Wong, Cara B. Ebbeling, David S. Ludwig, Brian A. Irving, Matthew C. Scott, James Stampley, Brett Davis, Neil Johannsen, Rachel Matthews, Cullen Vincellette, Andrea K. Garber, Gertraud Maskarinec, Ethan Weiss, Jennifer Rood, Alyssa N. Varanoske, Stefan M. Pasiakos, Steven B. Heymsfield, John A. Shepherd

## Abstract

**Background:** Recent three-dimensional optical (3DO) imaging advancements have provided more accessible, affordable, and self-operating opportunities for assessing body composition. 3DO is accurate and precise with respect to clinical measures made by dual-energy X-ray absorptiometry (DXA). However, the sensitivity for monitoring body composition change over time with 3DO body shape is unknown.

**Objective:** To evaluate 3DO’s ability to monitor body composition changes across multiple intervention studies.

**Methods:** A retrospective analysis was performed using intervention studies on healthy adults that were complimentary to the cross-sectional study, Shape Up! Adults. Each participant received a DXA (Hologic Discovery/A system) and 3DO (Fit3D ProScanner) scan at baseline and follow-up. 3DO meshes were digitally registered and reposed using Meshcapade to standardize the vertices and pose. Using an established statistical shape model, each 3DO mesh was transformed into principal components (PCs), which were used to predict whole-body and regional body composition values using published equations. Body composition changes (follow-up minus baseline) were compared to DXA with linear regression.

**Results:** The analysis included 133 participants (45 females) in six studies. The mean (SD) length of follow-up was 13 (5) weeks, range 3-23 weeks. Agreement between 3DO and DXA (R^2^) for changes in total fat mass (FM), total fat-free mass (FFM), and appendicular lean mass, respectively, were 0.86, 0.73, and 0.70 with RMSEs of 1.98 kg, 1.58 kg, and 0.37 kg in females, and 0.75, 0.75, and 0.52 with RMSEs of 2.31 kg, 1.77 kg, and 0.52 kg in males. Further adjustment with demographic descriptors improved the 3DO change agreement to changes observed with DXA.

**Conclusions:** As compared to DXA, 3DO was highly sensitive in detecting body shape changes over time. The 3DO method was sensitive enough to detect even small changes in body composition during intervention studies. The safety and accessibility of 3DO allows users to self-monitor on a frequent basis throughout interventions.

## INTRODUCTION

Obesity remains an area of concern as global prevalence continues to rise (1-3). According to the Center for Disease Control and Prevention (CDC), more than 42% of adults in the United States were considered obese in 2018, while the prevalence was approximately 30% two decades prior (4). To counter the obesity epidemic, diet and physical activity interventions have been studied extensively to target weight loss (5, 6). However, metanalyses have shown that weight is only loosely associated with metabolic health (7) and initial changes in response to intervention are small and quickly undone long-term (6, 8). On the other hand, a range of changes in body composition (reduced total body, abdominal and visceral fat, and increased muscle mass) can be produced through diet or exercise intervention and are consistently associated with decreased cardiovascular disease risk (9, 10). Further, decrements in skeletal muscle, particularly in the elderly, can lead to losses in strength and endurance, reductions in energy expenditure, and an increased risk of insulin resistance (11). Nevertheless, measures of body composition have been relegated to research and specialized facilities while clinical care continues to rely on weight as a flawed marker of health.

Another pressing reason to limit our reliance on weight is the differing relative weight of the compartments and tissues. Garrow (12) suggested, and Prentice et al. (13) concurred, that weight loss is typically 25% fat-free mass (FFM, i.e. lean soft tissue + bone mineral content) and 75% fat loss. However, recent research suggested this “25/75 rule of thumb” may not accurately describe various weight loss interventions. The amount FFM lost depends on energy intake, diet composition, sex, baseline adiposity, inactivity or type and physical activity level, and potentially metabolic and hormonal responses (14, 15). As such, it may not be appropriate to only monitor weight in interventions as FFM might be lost at a greater proportion. By monitoring body composition, investigators receive a more accurate assessment of their intervention’s efficacy.

Body composition by dual-energy X-ray absorptiometry (DXA) has been used extensively in clinical settings for its accuracy and precision of whole-body and regional measurements (16). Although DXA provides clinically useful measurements (e.g., bone mineral density, VAT, fat, and lean mass), it requires expensive radiological equipment, qualified technicians, and may not be feasible or accessible for routine clinical practice or frequent monitoring. The ideal body composition method should be affordable, accessible, free of ionizing radiation, and not require radiological-qualified technicians.

Three-dimensional optical (3DO) imaging has become increasingly accessible, made large advancements in recent years, and is safe to use repeatedly (17, 18). 3DO scanners provide accurate and precise digital anthropometry in comparison to criterion methods and output a 3D mesh that represents a person’s entire shape (19, 20). 3DO shape has shown to be highly predictive of DXA body composition (21-24). The next pressing issue is whether 3DO could successfully capture *changes* in body composition as different modalities have not validated well longitudinally to standard methods (25). However, the lack of available longitudinal data has limited the assessment of 3DO in monitoring body composition changes. The hypothesis of the study was that 3DO can monitor change with a similar sensitivity to DXA given previous cross-sectional accuracy and precision. Therefore, the objective of this study was to evaluate 3DO’s accuracy for monitoring body composition changes across a variety of intervention studies in comparison to DXA.

## METHODS

### Study Design

The current study was a retrospective analysis of six complimentary intervention studies to the Shape Up! Adults study (NIH R01 DK109008, ClinicalTrials.gov ID NCT03637855), which was originally a cross-sectional study with a planned longitudinal arm. In order to study 3DO’s ability to monitor body composition over time, collaborators adopted our DXA and 3DO protocol. The studies included Time-restricted Eating on Weight Loss (TREAT) (26); Macronutrients and Body Fat Accumulation: A Mechanistic Feeding Study (FB4: Framingham, Boston, Bloomington, Birmingham, and Baylor) (27-29); Resistance Exercise and Low-Intensity Physical Activity Breaks in Sedentary Time to Improve Muscle and Cardiometabolic Health Pilot Study (REALPA, NIH R21AG058181, ClinicalTrials.gov ID NCT03771417) (30), Louisiana State University (LSU) Athletes; Trial of Testosterone Undecanoate for Optimizing Performance During Military Operations (OPS II) (31, 32); and patients with bariatric surgery. If available, study-specific information (e.g., site, protocol, aims) can be found on ClinicalTrials.gov (**Table 1**). All study protocols were previously approved by their respective Institutional Review Board.

**Table 1.** Descriptions of longitudinal studies.

| **Study Name (Acronym)** | **Sex (N)** | **Intervention** | **Clinical Trials Number** |
| --- | --- | --- | --- |
| Athletes | Female 5 | Basketball team during training | NA |
| Bariatric | Female 2 | Bariatric surgery | NA |
| Macronutrients and Body Fat Accumulation: A Mechanistic Feeding Study (FB4: Framingham, Boston, Bloomington, Birmingham, and Baylor) | Female 15; Male 40 | Hypocaloric diet with low carbohydrate intake | NCT03394664 |
| Resistance Exercise and Low-Intensity Physical Activity Breaks in Sedentary Time to Improve Muscle and Cardiometabolic Health (REALPA) | Female 8; Male 5 | 16 weeks of resistance exercise with or without low intensity physical activity breaks in sedentary time or moderate intensity aerobic exercise in older adults | NCT03771417 |
| Time Restricted Eating on Weight Loss (TREAT) | Female 15; Male 27 | Time-restricted diet (16 hour fast, 8 hours to feed) | NCT03393195 |
| Trial of Testosterone Undecanoate for Optimizing Performance During Military Operations (OPS II) | Male 16 | Simulated military operational stress | NCT04120363 |

Complete study protocols can be found on https://clinicaltrials.gov/.

The TREAT participants were only allowed to consume food between 12 pm and 8 pm with the goal of fat loss (26). The FB4 cohort was given a hypocaloric, low-carbohydrate diet with the goal to lose 15±3% of baseline body weight (27-29). The REALPA study introduced whole-body resistance exercise (2 days/week) alone (i), or with moderate intensity aerobic exercise (50 min/day, 3 days/week at 4 METS) (ii), or low-intensity physical activity (LPA) breaks in sedentary time (∼10 min/break, 6 breaks/day, 5 days/week at 2METS) in adults 65-80 years of age to observe changes to muscle and cardiometabolic health markers after the 16-week long intervention. A subset of the REALPA participants who had both 3DO and DXA scans available for analysis were included in this retrospective study. (30). The LSU athletes were female basketball players that were measured at the beginning and through training camp (ranging 1-5 months), evaluating body composition as a result of a preseason training program. The OPS II study tested the effects of an intramuscular injection of testosterone undecanoate compared to placebo on changes in muscle and fat mass in recreationally-active young males undergoing simulated military operational stress consisting of sleep restriction, high exercise-induced energy expenditure, and limited energy intake (31, 32). The bariatric patients who were recruited from the University of California, San Francisco (UCSF) received bariatric surgery (surgery type was not recorded) with the goal of weight-loss. Although weight loss or body composition changes was not the goals of each study, the current aim of this analysis was to evaluate 3DO’s ability to monitor body composition changes in comparison to DXA.

### Participants

All participants provided informed consent before participation. Participants were deemed ineligible for this analysis if they were pregnant, breast feeding, had missing limbs, non-removable metal (e.g., joint replacements), previous body-altering surgery (e.g., breast augmentation), unable to stand still for one minute, or lie still for 3 minutes. Participants received same-day whole-body 3DO and DXA scans at baseline and follow-up. If the study had two follow-up appointments, the first of the two was used and will be considered as the “follow-up” in the current analysis. Participants were excluded from the analysis if they were missing either baseline or follow-up data from either DXA or 3DO.

### DXA

Height and weight were measured prior to the DXA scan. Participants received a single whole-body DXA scan with a Hologic Discovery/A system (Hologic Inc., Marlborough, Massachusetts) according to International Society for Clinical Densitometry guidelines (33). Participants laid supine on the scanning bed and were positioned by the DXA technician with arms by the side and feet internally rotated. Scans took approximately three minutes for the whole-body scan. All raw scans from each study were securely transferred to the University of Hawaii Cancer Center and analyzed by a single certified technologist using Hologic Apex version 5.6 with the National Health and Nutrition Examination Survey Body Composition Analysis calibration option disabled (34). After analysis, proprietary algorithms automatically generate body composition values. DXA measurements used in this analysis included whole-body and regional (i.e., arms, legs, trunk) fat and fat-free measures.

### Three-Dimensional Optical

Participants changed into form-fitting tights, a swim cap, and a sports bra if female. Female participants from the FB4 used form-fitting swimsuits. 3DO surface scans were taken on the Fit3D ProScanner version 4.x (Fit3D Inc., San Mateo, California). Participants grasped telescoping handles on the scanner platform and stood upright with shoulders relaxed and arms positioned straight and abducted from their torso. The platform rotates once around and takes approximately 45 seconds for the completion of the scan. Final point clouds were converted to a mesh connected by triangles with approximately 300,000 vertices and 600,000 faces representing the body shape (22).

Fit3D meshes were sent to Meshcapade (Meshcapade GmbH, Tubingen, Germany) for registration and to be digitally reposed. Their algorithm registers each mesh to a 110,000-vertex template with complete anatomical correspondence. Each vertex corresponds to a specific anatomical location across all registered meshes. All meshes were digitally reposed to a T-pose, where the person was standing straight, arms were brought horizontal and in the plane with the body, and arms and legs were straightened (35). The registered meshes were transformed into principal component (PC) space from an established statistical shape model (24). Principal component analysis orthogonalizes and reduces the dimensionality of the data so that fewer variables are needed to describe the data’s variance (21). Total FM and regional (i.e., arms, legs, trunk, VAT) body composition estimates were derived previously using either exclusively PC descriptors of shape or PC descriptors with demographic adjustments (24). In this study, PC only body composition equations were used since complete demographics and anthropometrics were not available on all cohorts. Total FFM and percent fat (%fat) were derived dependently from total FM and total body mass. Appendicular lean mass (ALM) was defined as the sum of lean soft tissue masses for legs and arms by convention (36). ALM index (ALMI) was derived by dividing ALM by height-squared.

Average 3DO body shape representations were created to visualize the average shape at baseline and follow-up for each intervention study. PCs were averaged by study intervention to make an average vector of PCs, which was then inverted back into coordinate space (x, y, and z) to acquire the image.

### Statistical Analysis

Body composition change for all variables was defined as follow-up minus baseline. 3DO body composition change was compared to DXA body composition change using linear regression. Bland-Altman plots were made for the cross-sectional comparisons. Coefficient of determination (R^2^), Lin’s concordance coefficient (CCC), and root mean square error (RMSE) were used to report the relationship and accuracy of the comparison. Scatter plots were used to visualize the comparison. Additional adjustments were made to explain any potential bias using stepwise forward linear regression and five-fold cross-validation. Potential covariates included ethnicity and baseline height, weight, BMI, and age as well as changes in weight, height, and BMI. Covariates remained in the model if they had a p-value <0.05. The least significant change (LSC) was used to determine if the change in the body composition measure was significantly different (95% confidence) than zero (37). Student’s t-test was used to evaluate mean differences between 3DO and DXA outputs. P-value < 0.05 was considered statistically significant.

The LSCs used in this analysis were derived from Wong et al. (24). LSC is defined as 2.77 x precision error (21, 37). Female LSC for FM, FFM, and %fat was 1.52 kg, 1.52 kg, and 2.27% for 3DO, while 0.64 kg, 0.75 kg, and 0.91% for DXA, respectively. Male LSC for FM, FFM, and %fat was 1.22 kg, 1.22 kg, and 1.58% for 3DO, while 0.69 kg, 0.94 kg, and 0.78% for DXA, respectively. Since ALM was not reported in the previous publication, in order to get the precision error to calculate the LSC, the test-retest precision for ALM was derived using the same sample from Wong et al. (24). Lastly, Cohen’s kappa analysis was utilized to assess the consistency between the DXA and 3DO report for each level, considering any agreement that may have happened due to chance. The kappa scores can be interpreted as follows: 0-0.20 = no agreement, 0.21-0.39 = fair agreement, 0.40-0.59 = moderate agreement, 0.60-0.79 = substantial agreement, 0.80-0.99 = near perfect agreement, and 1 = perfect agreement (38).

To test Garrow’s (12) and Prentice et al.’s (13) suggestion that 25% of weight loss is FFM, change in total FFM was divided by the absolute change in weight multiplied by 100 ([ΔFFM /absolute Δweight]*100). This was applied to those who had a negative weight change (weight loss). The absolute change in weight was used in order to evaluate gains or losses in FFM.

Experimental models were created using the change in PCs (Δ PCs) to test if different modeling methods would improve the body composition change predictions. Model 1 used the Δ PCs, model 2 used the Δ PCs and baseline total fat mass, model 3 used the Δ PCs and baseline PCs, model 4 used the baseline and follow-up PCs, and model 5 used the Δ PCs and change in weight. These models were built with stepwise forward linear regression with five-fold cross-validation. All statistical analyses were done in R version 4.2.1 (R Core Teams, Vienna, Austria).

## RESULTS

One hundred and thirty-three participants (43 female and 85 male) were included in the final analysis (**Table 2**). One-hundred sixty-four participants were excluded for dropout (n=4), unavailable 3DO or DXA data at one or both timepoints (n=157), movement artifacts (n=2), or mislabeled data (n=1) (**Supplemental Figure 1**). The time between baseline and follow-up DXA scans across all studies ranged from three to twenty-three weeks (**Figure 1**). Females and males lost on average 3.5 kg and 5.4 kg of total FM, 1.8 kg and 3.4 kg of total FFM, and 30 g and 100 g of VAT as a result of interventions, respectively, according to DXA. The majority of the body composition changes occurred in the trunk for both sexes. Average body shapes were presented at baseline and follow-up for each study intervention (**Figure 2**).

**Table 2.** Sample characteristics at baseline and follow-up visits

|  | Female<br>(N=45) |  | Male<br>(N=88) |  |
| --- | --- | --- | --- | --- |
| **Age (years)** | **Baseline** | **Follow-up** | **Baseline** | **Follow-up** |
| Mean (SD) | 45.6 (15.7) | 45.8 (15.7) | 36.3 (13.3) | 36.5 (13.3) |
| Median [Min, Max] | 45.9 [18.5, 76.7] | 46.2 [18.8, 77.1] | 32.0 [19.0, 73.5] | 32.0 [19.3, 73.9] |
| **Ethnicity** |  |  |  |  |
| Asian | 3 (6.7%) |  | 11 (12.4%) |  |
| Black | 10 (22.2%) |  | 7 (7.9%) |  |
| Hispanic | 5 (11.1%) |  | 7 (7.9%) |  |
| White | 27 (60.0%) |  | 64 (71.9%) |  |
| **Height (cm)** |  |  |  |  |
| Mean (SD) | 165 (7.46) |  | 177 (6.67) |  |
| Median [Min, Max] | 164 [149, 185] |  | 176 [162, 194] |  |
| **Weight (kg)** |  |  |  |  |
| Mean (SD) | 86.7 (17.5) | 81.4 (13.2) | 97.6 (17.1) | 89.4 (17.3) |
| Median [Min, Max] | 82.3 [63.5, 143] | 80.6 [63.0, 116] | 95.2 [61.5, 142] | 87.3 [58.9, 175] |
| **BMI (kg/m<sup>2</sup>)** |  |  |  |  |
| Mean (SD) | 31.8 (6.61) | 29.9 (5.00) | 31.3 (5.42) | 28.7 (5.54) |
| Median [Min, Max] | 30.4 [21.8, 51.3] | 29.5 [21.7, 43.7] | 30.1 [19.8, 45.8] | 27.9 [19.0, 57.3] |
| **DXA Total FM (kg)** |  |  |  |  |
| Mean (SD) | 34.8 (11.4) | 31.3 (8.80) | 28.6 (10.8) | 23.2 (9.34) |
| Median [Min, Max] | 34.6 [12.9, 68.4] | 31.0 [14.0, 54.3] | 28.6 [9.14, 67.3] | 22.6 [6.00, 53.8] |
| **DXA Total FFM (kg)** |  |  |  |  |
| Mean (SD) | 52.3 (8.35) | 50.5 (7.03) | 69.6 (9.24) | 66.2 (8.37) |
| Median [Min, Max] | 52.4 [37.5, 77.0] | 49.7 [35.9, 67.8] | 68.0 [51.4, 91.6] | 65.4 [49.6, 90.9] |
| **DXA Percent Fat (%)** |  |  |  |  |
| Mean (SD) | 39.3 (7.21) | 37.8 (6.66) | 28.3 (6.96) | 25.2 (7.07) |
| Median [Min, Max] | 41.8 [19.9, 51.5] | 40.2 [20.6, 46.9] | 29.1 [11.1, 47.4] | 25.9 [7.81, 43.9] |
| **DXA ALM (kg)** |  |  |  |  |
| Mean (SD) | 11.3 (2.17) | 11.0 (1.87) | 15.8 (2.23) | 15.1 (2.04) |
| Median [Min, Max] | 11.6 [7.31, 15.6] | 10.7 [7.52, 15.0] | 15.8 [11.2, 20.9] | 14.8 [10.8, 20.9] |
| **DXA VAT (kg)** |  |  |  |  |
| Mean (SD) | 0.56 (0.27) | 0.53 (0.26) | 0.56 (0.28) | 0.46 (0.23) |
| Median [Min, Max] | 0.51 [0.13, 1.10] | 0.52 [0.12, 1.29] | 0.50 [0.16, 1.40] | 0.40 [0.13, 1.15] |
| **DXA Arm FM (kg)** |  |  |  |  |
| Mean (SD) | 2.14 (0.83) | 1.92 (0.60) | 1.78 (0.84) | 1.46 (0.67) |
| Median [Min, Max] | 2.04 [0.73, 5.32] | 1.94 [0.79, 3.53] | 1.61 [0.52, 4.94] | 1.39 [0.36, 3.82] |
| **DXA Arm FFM (kg)** |  |  |  |  |
| Mean (SD) | 2.68 (0.51) | 2.59 (0.44) | 4.34 (0.73) | 4.13 (0.66) |
| Median [Min, Max] | 2.62 [1.79, 4.01] | 2.49 [1.80, 3.74] | 4.30 [2.93, 6.42] | 4.10 [2.78, 6.19] |
| **DXA Leg FM (kg)** |  |  |  |  |
| Mean (SD) | 6.54 (2.18) | 5.97 (1.75) | 4.67 (1.70) | 3.86 (1.42) |
| Median [Min, Max] | 6.02 [2.90, 13.1] | 5.89 [3.13, 10.8] | 4.45 [1.65, 10.9] | 3.88 [1.08, 9.16] |
| **DXA Leg FFM (kg)** |  |  |  |  |
| Mean (SD) | 8.64 (1.72) | 8.37 (1.48) | 11.4 (1.61) | 11.0 (1.48) |
| Median [Min, Max] | 8.75 [5.52, 12.0] | 8.24 [5.72, 11.3] | 11.3 [7.94, 15.1] | 10.8 [7.62, 15.1] |
| **DXA Trunk FM (kg)** |  |  |  |  |
| Mean (SD) | 16.3 (6.16) | 14.4 (4.79) | 14.4 (6.10) | 11.3 (5.48) |
| Median [Min, Max] | 16.6 [4.61, 34.9] | 14.8 [4.73, 24.5] | 14.1 [3.52, 34.3] | 10.8 [2.16, 26.6] |
| **DXA Trunk FFM (kg)** |  |  |  |  |
| Mean (SD) | 26.1 (4.31) | 25.0 (3.81) | 33.9 (4.88) | 31.9 (4.44) |
| Median [Min, Max] | 25.7 [19.2, 41.6] | 23.9 [17.5, 37.4] | 33.5 [25.1, 46.4] | 31.7 [23.8, 44.3] |

Abbreviations: FM (fat mass); FFM (fat-free mass); ALM (appendicular lean mass); DXA (dual-energy X-ray absorptiometry); VAT (visceral adipose tissue)

**Figure 1.**
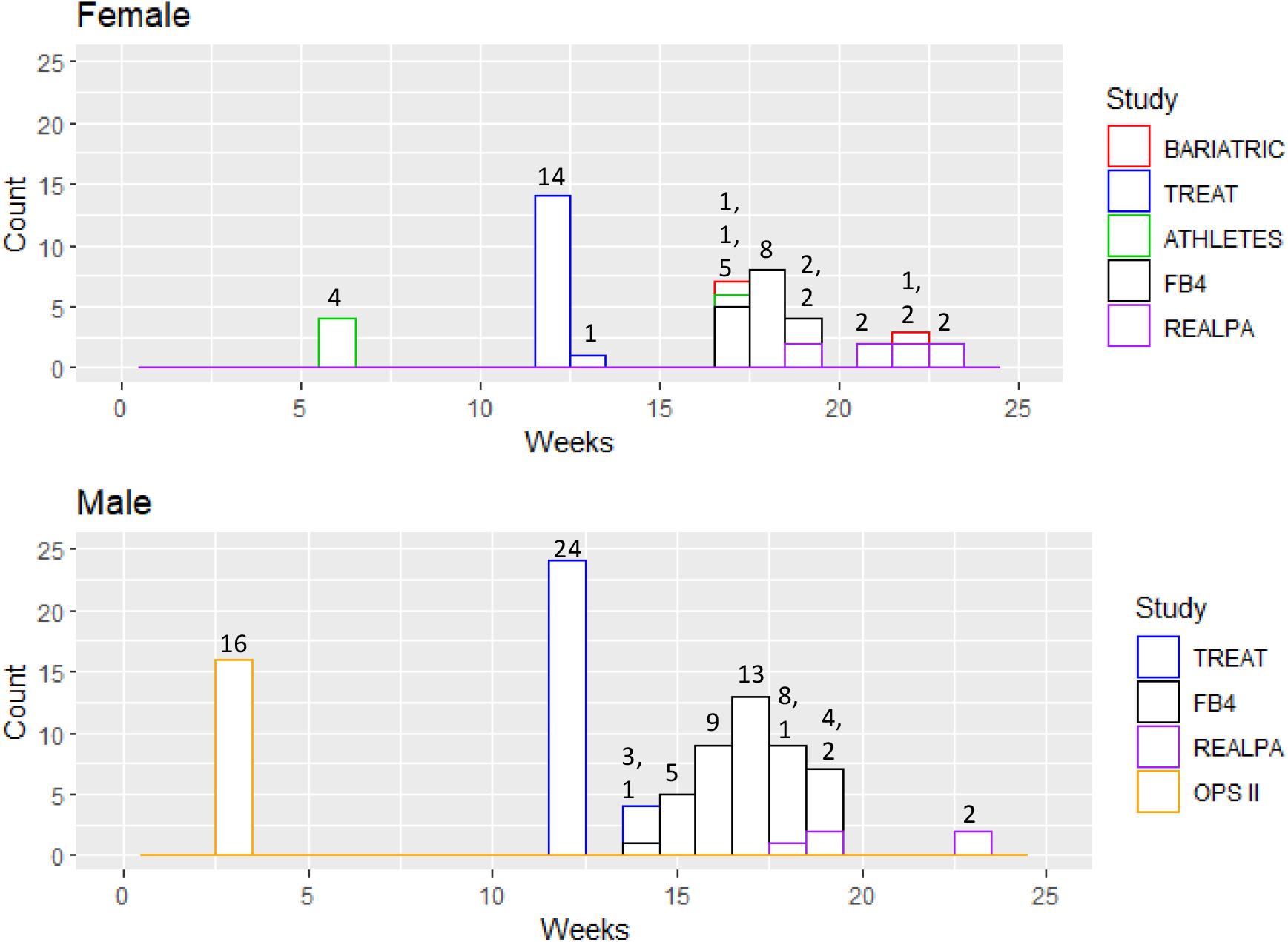
Histograms of the time (weeks) between the baseline and follow-up DXA scans for females (top) and males (bottom). The numbers over the bars represent the count. Multiple numbers over a multi-color bar represent the count in the corresponding order.

**Figure 2.**
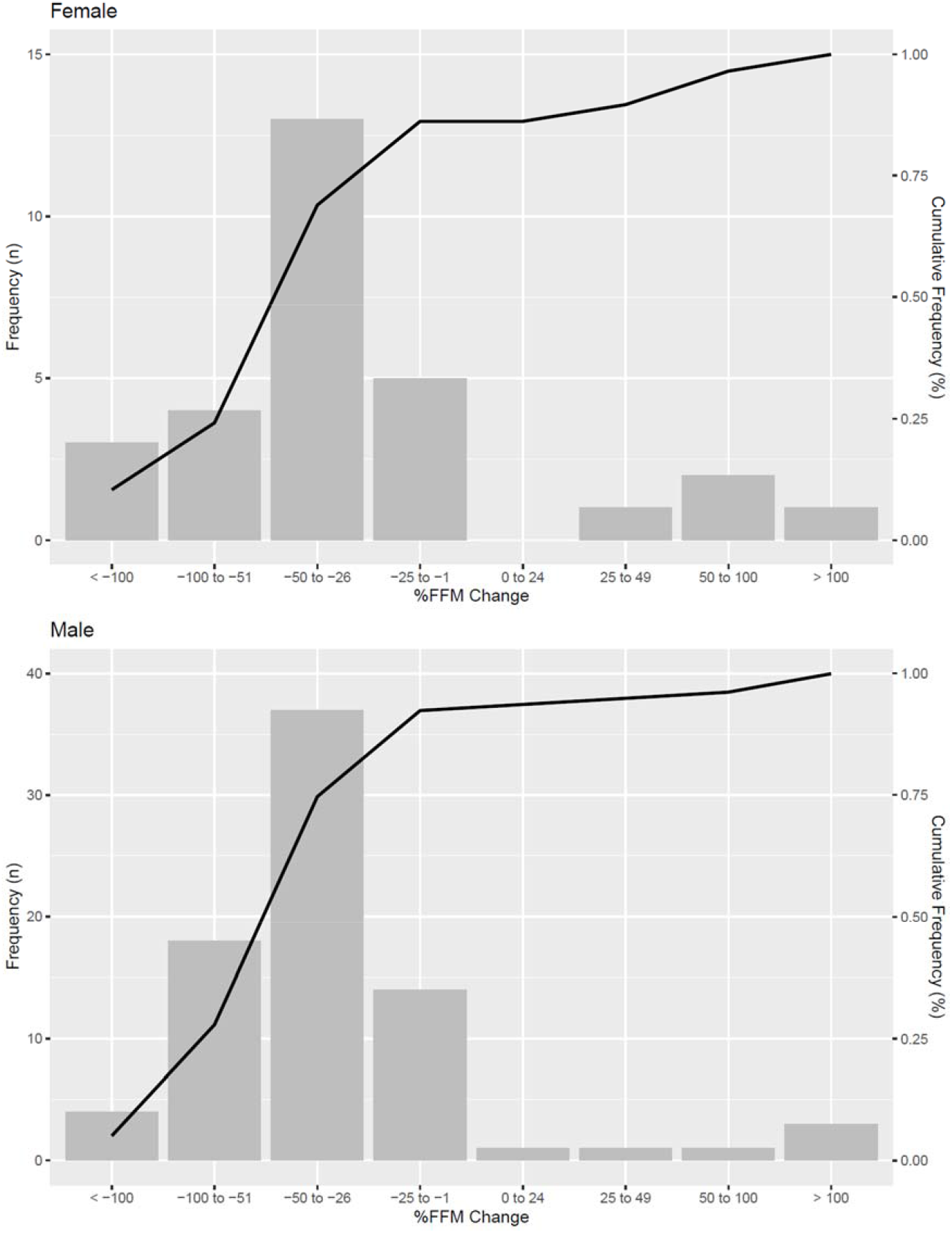
Average baseline (left) and follow-up (right) body shapes for females (top) and males (bottom) by study.

In females (**Figure 3**), 3DO and DXA total FM and FFM at baseline (R^2^s; 0.91 and 0.84; RMSE; 3.3 kg and 3.3 kg, respectively) and follow-up (R^2^s; 0.89 and 0.84; RMSE; 2.8 kg and 2.8 kg, respectively) were highly correlated, while %fat and ALM were moderately correlated (R^2^s: 0.61 – 0.79). Female Bland-Altman plots are shown in **Supplemental Figure 2**. Slight proportional bias was observed in the baseline total FM from a single outlier with high leverage but not seen in the follow-up. In the comparison of 3DO body composition changes to DXA (Supplemental Table 1), 3DO achieved an R^2^ of 0.86, 0.73, 0.23; and 0.70, CCC of 0.90, 0.82, 0.47, and 0.81; RMSE of 1.98 kg, 1.58 kg, 2.2%, 0.37 kg for total FM, total FFM, %fat, and ALM, respectively. Mean differences were observed for VAT, ALM, ALMI, and leg FFM (30 g, 0.19 kg, 0.07 kg/m^2^, 0.16 kg, respectively, p-value < 0.05). After adjustments for possible covariates in the stepwise linear regression models with forward selection (**Supplemental Table 2**), weight change further explained variance in total FM, FFM, and ALM, while changes in BMI further explained variance in %fat change, which modestly improved the R^2^s to 0.90, 0.80, 0.51, and 0.79, respectively. The adjustments with demographic covariates alleviated residual bias between 3DO and DXA change. Mean differences (i.e., VAT, ALM, ALMI, and leg FFM) were no longer observed after adjustments (p-value = 0.99). The green glyphs (**Figure 3 and 4**) symbolizes the participants that exceeded both the DXA and 3DO LSCs, the purple glyphs were those who did not exceed DXA nor 3DO’s LSC, and the orange and blue glyphs exceeded one LSC but not the other. The LSCs were depicted with the orange vertical lines (3DO) or the blue horizontal lines (DXA).

**Figure 3.**
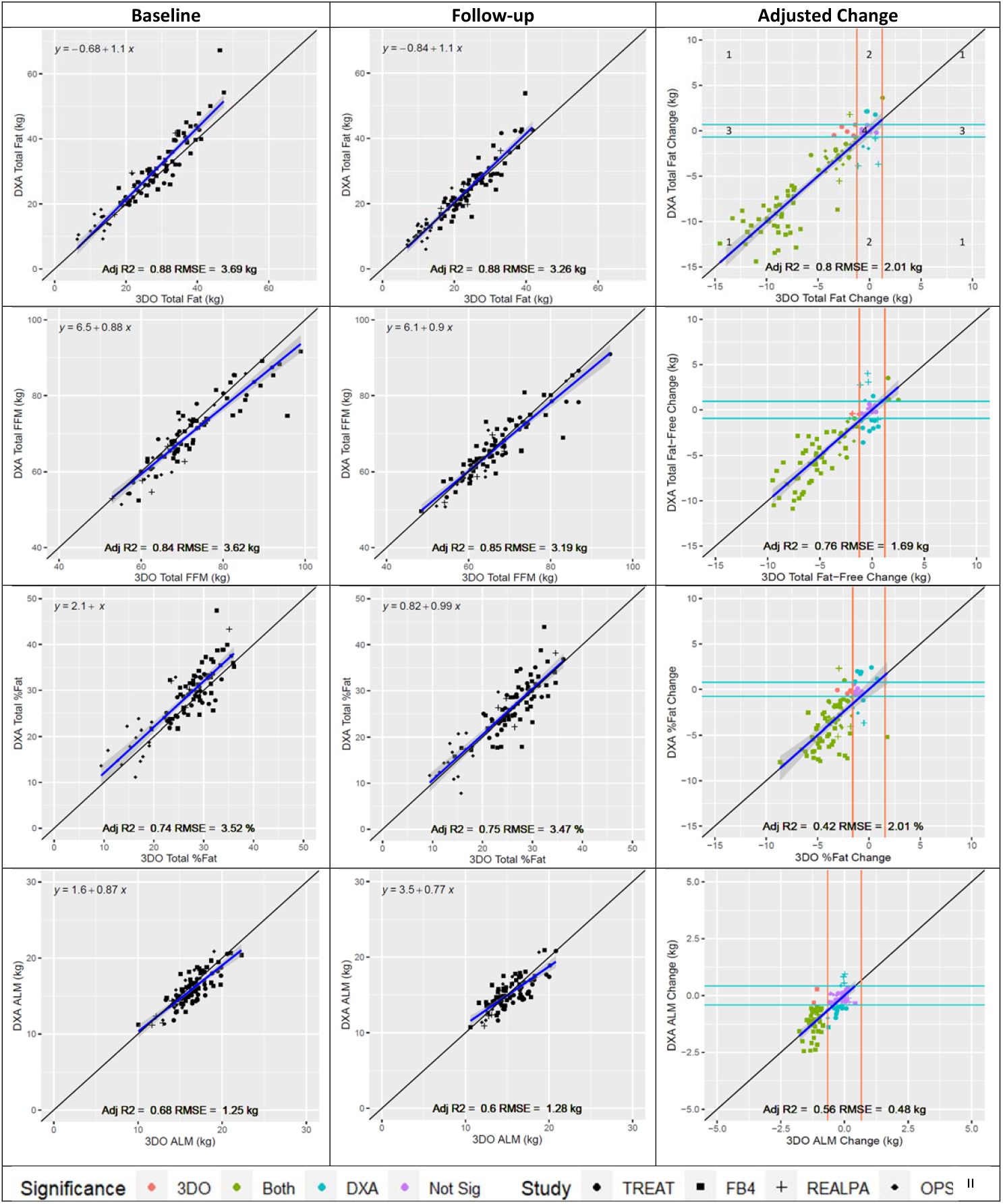
(Female) Scatter plot comparisons between 3DO and DXA body composition at baseline, follow-up, and adjusted change. Blue, horizontal lines and orange, vertical lines are signifying the amount of change needed to pass the least significant change for DXA and 3DO, respectively. Purple (Zone 4) and green glyphs (Zone 1) represent agreement of significant or nonsignificant change by 3DO and DXA. The orange glyphs (Zone 3) represent significant change detected by 3DO but not DXA and vice versa for the blue glyphs (Zone 2). The zones and glyphs are consistent for the proceeding adjusted change plots.

**Figure 4.**
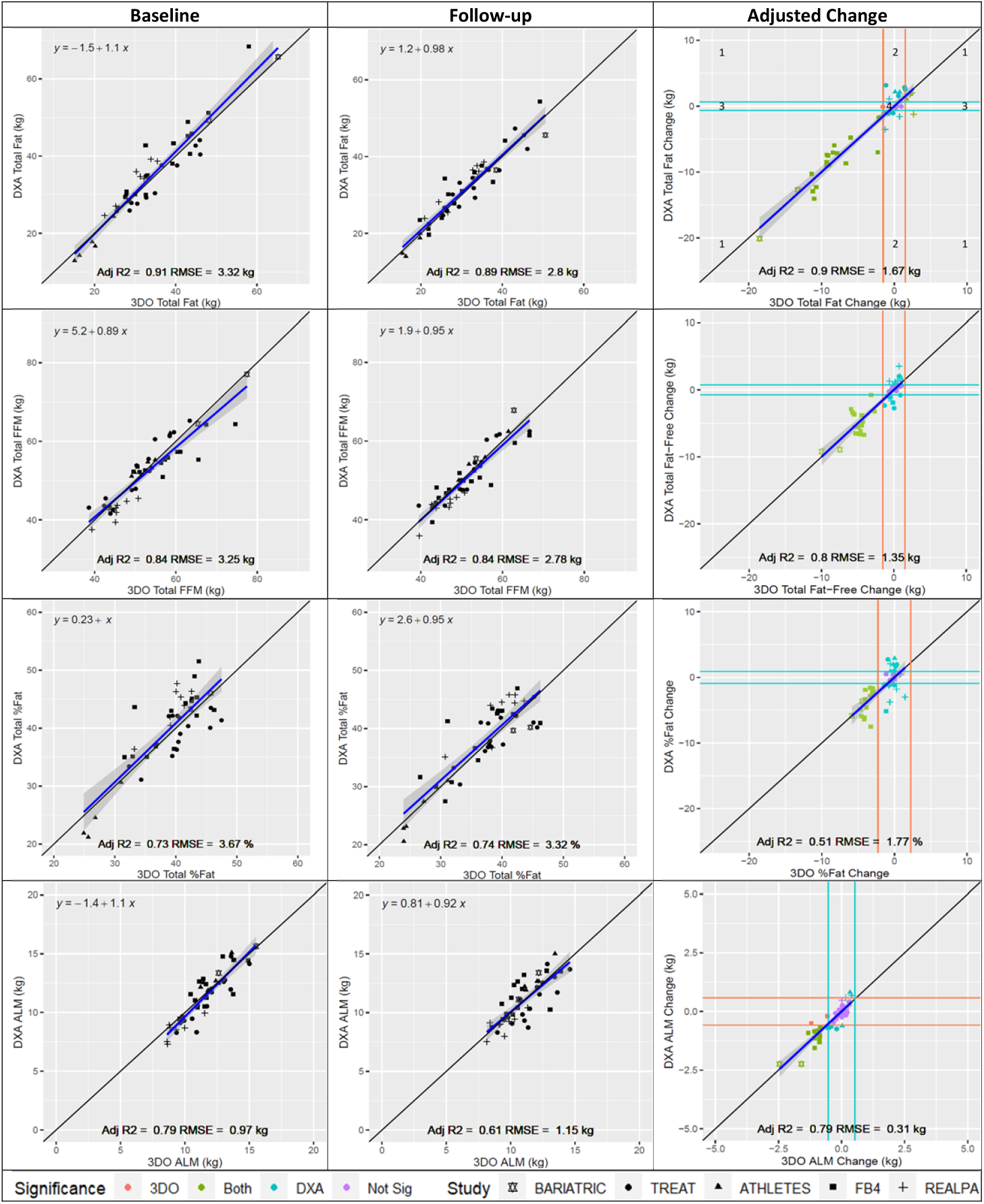
(Male) Scatter plot comparisons between 3DO and DXA body composition at baseline, follow-up, and adjusted change. Blue, horizontal lines and orange, vertical lines are signifying the amount of change needed to pass the least significant change for DXA and 3DO, respectively. Purple (Zone 4) and green glyphs (Zone 1) represent agreement of significant or nonsignificant change by 3DO and DXA. The orange glyphs (Zone 3) represent significant change detected by 3DO but not DXA and vice versa for the blue glyphs (Zone 2). The zones and glyphs are consistent for the proceeding adjusted change plots.

In males (**Figure 4**), 3DO and DXA total FM and FFM at baseline (R^2^s; 0.88 and 0.84; RMSE; 3.7 kg and 3.6 kg, respectively) and follow-up (R^2^s; 0.88 and 0.85; RMSE; 3.3 kg and 3.2 kg, respectively) were highly correlated, while %fat and ALM were moderately correlated (R^2^s: 0.60 – 0.75). Male Bland-Altman plots are shown in Supplemental Figure 3. Proportional bias was observed in total FM and %fat due to a single outlier with high leverage. Not bias was observed in total FFM and ALM. In the comparison of 3DO body composition changes to DXA (**Supplemental Table 1**), 3DO achieved an R^2^ of 0.75, 0.75, 0.25, and 0.52; CCC of 0.76, 0.78, 0.39, and 0.64; and RMSE of 2.3 kg, 1.8 kg, 2.4%, and 0.5 kg for total FM, total FFM, %fat, and ALM, respectively. Of the observed variables in **Figure 4**, mean differences were observed for total FM, total FFM, %fat, and ALM (p-value<0.001). After adjustments for possible covariates in stepwise linear regression with forward selection (**Supplemental Table 2**), R^2^s modestly improved for changes in total FM, FFM, %fat, and ALM to 0.80, 0.76, 0.42, and 0.56, respectively. Mean differences in total FM, total FFM, %fat, and ALM were no longer observed after adjustments (p-value = 0.99).

3DO predicted regional body composition changes (i.e., arms, legs, and trunk) with R^2^s that ranged from 0.39 – 0.91 and RMSEs that ranged from 0.16 – 1.3 kg in females and males (**Supplemental Table 1**). 3DO and DXA found significant changes in the majority of the sample for total FM (69%), total FFM (78%), %fat (53%), and ALM (78%) (**Supplemental Table 3**). Cohen’s kappa showed that 3DO had a moderate agreement to DXA for most outputs.

From the total sample, 64% of females and 90% of males lost weight after intervention. Of those that lost weight, % FFM loss relative to weight loss ranged from -1370% to 478% as measured by DXA. Among those who lost FFM, 80% of females and 81% of males lost more than 25% FFM relative to their weight loss (**Figure 5**). The % FFM loss relative to weight loss according to study was on average 36% for Bariatric surgery, 37% for FB4, 27% for OPS II, 313% for Athletes, 85% for TREAT, and 59% for REALPA. The very high percentages were for small weight changes and/or large compartment changes with little overall weight change (e.g., Δ FFM /absolute(Δweight) * 100 for an Athlete = (−1.96/0.36) * 100 = -544%).

**Figure 5.**
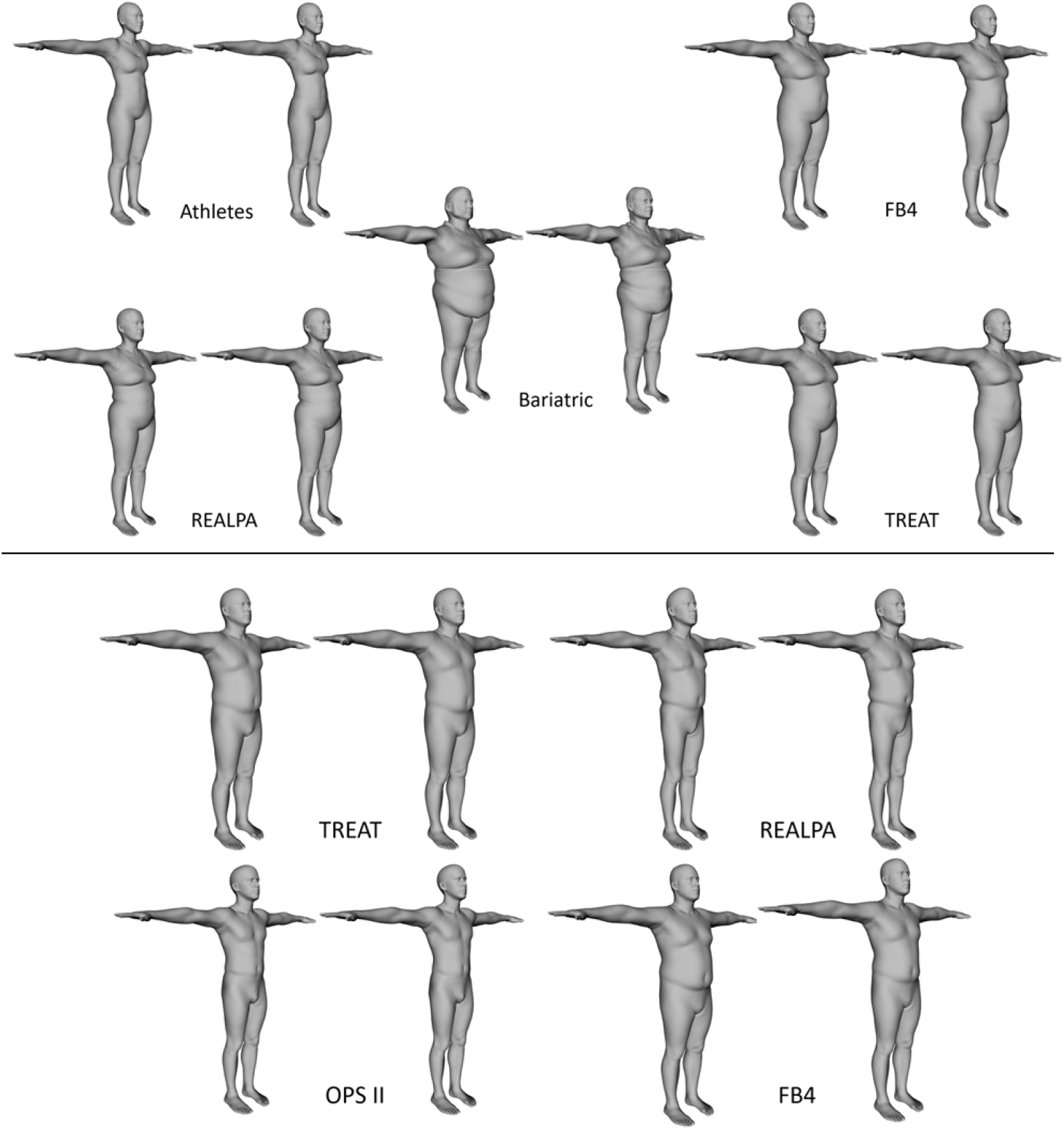
Histograms to show the frequency of percent fat-free mass change relative to total weight change in the female (top) and male (bottom) samples.

The experimental models (**Supplemental Tables 4 and 5**) for females (models 1-5) and males (models 1-3 and 5) predicted change better than the unadjusted results presented in **Supplemental Table 1**. Female models 1, 2, and 3 were the same as male models 1, 2, and 3 after the stepwise regression. The prediction of total FM change improved in both females (R^2^ = 0.94, RMSE = 1.29 kg) and males (R^2^ = 0.84, RMSE = 1.79 kg).

## DISCUSSION

These present findings support 3DO as a sensitive tool assessing changes in body composition, compared to DXA. The current study evaluated 3DO’s ability to monitor body composition change with body shape via the 3DO mesh. Previously derived shape models and equations were used to estimate body composition values (24), and 3DO change from baseline to follow-up was compared to DXA change. Overall, 3DO change was highly correlated to DXA change, and the two methods agreed on the statistical significance of the change in the majority of the study population. Additional adjustments of demographics explained further variance between 3DO and DXA change. The experimental models showed that specific calibration with changes in shape might improve the body composition change prediction. Additional longitudinal data may further validate the experimental models. Nevertheless, the cross-sectional 3DO models produced comparable body composition estimates compared to DXA, which supports our hypothesis that using 3DO imaging is a feasible method to monitor body composition changes.

With respect to the “rule-of-thumb” by Garrow’s (12) and Prentice et al. (13) that 25% of weight loss comes from FFM, the proportion of FFM lost by the vast majority of our sample was greater than 25% (**Figure 5**). Thus, this “rule-of-thumb” underestimated the loss of FFM for most participants. The loss of FFM varied by study intervention due to differences in protocol, type of activity, and objectives. This is a critical area of study given that loss of FFM is associated with diminished strength and decrements in physical functioning, exercise endurance and capacity, metabolic rate, and health (11). This further supports the idea that the relative proportion of FM and FFM change is dependent on energy intake, diet composition, sex, baseline adiposity, inactivity or type and level of physical activity level, and potentially metabolic and hormonal responses (14). It should also be noted that the experimental group in the OPS II study received testosterone, which has shown to improve muscle protein kinetics during stress and was a contributing factor to the proportion of FFM loss (32). The LSU athletes and OPS II participants were study examples that monitored body composition for performance. 3DO can also provide an accessible and safe method to monitor body composition changes since muscle build-up can play a pivotal role in strength, endurance, and speed. Some participants had percent changes in FFM that were greater than 100%. This was likely due to an undesirable recomposition where the person increased FM and lost FFM, which resulted in a minimal weight change (small denominator). Hydration status may have impacted individuals with minimal changes (15). Because DXA does not measure hydration status, DXA has assumptions of hydration built-in, while 3DO estimates were regressed to DXA. The current analysis further emphasized the importance of monitoring body composition, in addition to weight, to better understand changes in health status and performance factors.

In the current study, the change in 3DO FM was underestimated while FFM was overestimated in both females and males. These changes were not statistically significant in females but were significant in males. A potential reason for the male discrepancies could be that the current sample was beyond the adiposity range of the data used to train the original 3DO body composition models. The maximum percent fat of the training dataset was 38% (24), while the current population’s maximum was 47%. The training dataset lacked the shape variance of males with extreme adiposity and underestimated FM and %fat in this study. However, after adjusting for covariates, these discrepancies were no longer statistically significant. Using either the 3DO changes models with demographics or retraining the model with high adiposity participants could potentially address this issue.

Test-retest precision, also known as short-term precision, has an error that can be used to derive the LSC (2.77 x precision error) (37). Metrics with changes that surpass their LSC values were considered to be significantly greater than zero changes, with a 95% confidence. 3DO and DXA precision errors used for this study were taken from the Shape Up Adults group (24). To reduce the 3DO precision error, to be closer to that of DXA, and track body composition changes at the same significance, multiple 3DO scans could be taken and averaged at each time point. For total FM, the 3DO precision errors were 0.44 kg and 0.55 kg as well as 0.25 kg and 0.23 kg for DXA in males and females, respectively (24). Assuming a normal distribution of the precision errors, the precision of the averaged 3DO measure from multiple scans improves by the square root of the number of scans (e.g., the average of four scans would lower the precision error from 0.44 kg and 0.55 kg to 0.22 kg and 0.27 kg) (39). Unlike DXA, taking multiple scans with 3DO is fast and without additional radiation concerns. Dizziness can be a potential hazard, which is listed on the manufacturer warnings and our study consent form. However, our participants did not report dizziness even with multiple scans.

3DO VAT change compared to DXA VAT change had a low correlation, but this is likely to due to the compressed range of VAT change in our sample. In previous literature, DXA VAT has been shown to be highly correlated to gold standard, magnetic resonance imaging (MRI), cross-sectionally (R^2^ range: 0.81 – 0.86) (40-42). However, DXA VAT often has proportional bias for individuals with high VAT and has not validated well longitudinally compared to MRI (42, 43). Some of the bias may be explained by technique differences as MRI and CT are analyzed by a cross-section of the abdomen, while DXA is estimated from an X-ray’s 2D area. Nevertheless, given the cross-sectional correlation of DXA VAT to MRI and cardiometabolic markers, DXA VAT can still be considered an accessible tool to characterize health risks that can prompt health initiatives (44, 45). Since 3DO body shape has been shown to be correlated and predictive of DXA VAT and cardiometabolic markers (21), 3DO may be considered an accessible tool that can be used frequently due to its lack of ionizing radiation concerns. However, users should be wary of DXA and 3DO’s ability to accurately monitor VAT changes given the limited amount of validation in the literature.

The 3DO vs DXA RMSEs for comparing baseline or follow-up scans were higher (worse) than the RMSEs for the change comparisons (**Figures 3 and 4**). This may be due to the individual systematic bias expressed in the comparison of one technique (3DO) to another (DXA) that is subtracted away when looking at changes in the measures, and the fact that the precision error of both techniques was much lower than this individual systemic bias found in inter-technique comparisons of measures (46).

According to the authors’ knowledge, one other study has reported 3DO’s ability to monitor body composition change. Tinsley et al. (25) compared the changes in proprietary body composition estimates from 3DO scanners (Fit3D ProScanner, Size Stream SS20 (Size Stream LLC, Cary, NC, USA), and Styku S100 (Styku Comp. Los Angeles, CA, USA)) with changes in 4-compartment (4C) model measures in 21 volunteers. Changes in 3DO FM underperformed (CCC, 0.22 to 0.40 for total FM) compared to the 4C model. CCC for total FM in this study was much higher (CCC; 0.91 and 0.79) for females and males, respectively, showing better agreement with DXA. However, there were major differences between the two studies. This study had a larger sample size (n=128), greater ethnic-racial diversity, larger body composition changes, publicly available equations, and a different body composition modality (DXA instead of the 4C model, given that data for the 4C model was not available). Larger studies are warranted to fully evaluate 3DO’s ability to monitor change using the scanners’ proprietary body composition outputs.

This study had several strengths including sample size, variety and lengths of interventions, and an ethnically diverse sample. A study with 128 participants is currently one of the larger body composition change studies. Although the majority of the participants were ethnically white, there was still a representation of other ethnic backgrounds. This study included a variety of diet, physical activity, and surgical interventions that ranged from 3 to 23 weeks and included participants with large as well as little to no body composition changes.

This study is not without limitations. Although the sample size was rather large for a longitudinal analysis, a representative test set could not be made to validate the adjusted and experimental linear models. In addition, results from this study may not be generalizable to populations with poor health status or children (< 18 years old). Future work would be required to address gaps in knowledge in different populations such as infants and children, the accuracy of change in specific subgroups (i.e., BMI, ethnicity, and age), and the assessment of more accessible 3DO technology.

## CONCLUSION

This study indicated that 3DO body composition changes were highly correlated to DXA and show good feasibility to monitor changes over a variety of interventions. As the accessibility and popularity of 3DO continues to grow, more people will be able to use this technology to monitor their body composition in clinical and nonclinical settings. The findings of this study extend the 3DO literature, which has been limited to cross-sectional performance.

## Data Availability

All data produced in the present study are available upon reasonable request to the authors.

https://shepherdresearchlab.org/request-data/

## Abbreviations

3DO: (three-dimensional optical)
ALM: (appendicular lean mass)
ALMI: (appendicular lean mass index)
CV: (coefficient of variation)
DXA: (dual-energy X-ray absorptiometry)
FM: (fat mass)
FFM: (fat-free mass)
LSC: (least significant change)
PCA: (principal component analysis)
PCs: (principal components)
VAT: (visceral adipose tissue)

## Acknowledgements

We thank all the participants for graciously giving their time to be part of the studies, our collaborators at each site for their part in the study, Tyler Carter and Greg Moore at Fit3D for providing us the 3DO data, and Naureen Mahmood and Talha Zaman for providing us the application program interface to register and repose our 3DO data. The data underlying this study cannot be made publicly available because the data contains patient identifying information. Data is available from the Shape Up! Studies for researchers who meet the criteria for access to confidential data. For details and to request an application, please contact John Shepherd or visit www.shapeup.shepherdresearchlab.org.

## Author Contributions

MCW and JAS designed and conducted the research; MCW, LL, JB, IT, YEL, GM, and JAS were part of the data analysis; JMWW, NNK, CBE, BI, MCS, JS, BD, NMJ, RM, CV, DSL, EW, SBH, and JAS were in charge of their respective study recruitment and protocols; MCW and JAS drafted the manuscript and were responsible over the final content. All authors reviewed and approved the final manuscript.

