## Supplement for "Monitoring Body Composition Change for Intervention Studies with Advancing 3D Optical Imaging Technology in Comparison to Dual-Energy X-Ray Absorptiometry"

Supplemental Table 1. Unadjusted comparison of the change in 3DO to change in DXA whole-body and regional body compositions with linear regression

|  | **Female** | | | | **Male** | | | |
| --- | --- | --- | --- | --- | --- | --- | --- | --- |
|  | **R^2^** | **CCC** | **RMSE** | **Mean Difference (3DO-DXA)** | **R^2^** | **CCC** | **RMSE** | **Mean Difference (3DO-DXA)** |
| **Total FM** | 0.86 | 0.90 | 1.98 | -0.16 | 0.75 | 0.76 | 2.31 | **-1.54** |
| **Total FFM** | 0.73 | 0.82 | 1.58 | 0.16 | 0.75 | 0.78 | 1.77 | **1.54** |
| **Percent Fat** | 0.23 | 0.47 | 2.24 | -0.01 | 0.25 | 0.39 | 2.36 | **-1.53** |
| **VAT** | 0.31 | 0.53 | 0.09 | **0.03** | 0.23 | 0.47 | 0.11 | -0.01 |
| **ALM** | 0.70 | 0.81 | 0.37 | **0.19** | 0.52 | 0.64 | 0.52 | **0.39** |
| **ALMI** | 0.71 | 0.80 | 0.13 | **0.07** | 0.51 | 0.63 | 0.17 | **0.12** |
| **Arm FM** | 0.71 | 0.83 | 0.21 | 0.01 | 0.56 | 0.66 | 0.22 | **-0.08** |
| **Arm FFM** | 0.39 | 0.63 | 0.16 | 0.03 | 0.40 | 0.63 | 0.21 | 0.03 |
| **Leg FM** | 0.76 | 0.85 | 0.42 | 0.10 | 0.56 | 0.63 | 0.47 | **-0.31** |
| **Leg FFM** | 0.71 | 0.80 | 0.27 | **0.16** | 0.41 | 0.54 | 0.43 | **0.36** |
| **Trunk FM** | 0.91 | 0.92 | 0.91 | -0.31 | 0.77 | 0.79 | 1.31 | **-0.74** |
| **Trunk FFM** | 0.54 | 0.74 | 1.17 | -0.01 | 0.66 | 0.81 | 1.23 | 0.19 |

Abbreviations: ALM (appendicular lean mass); ALMI (appendicular lean mass index), CCC (Lin’s Concordance Coefficient), FM (fat mass); FFM (fat-free mass); VAT (visceral adipose tissue);

The outcome variables are from DXA, and predictor variables are from 3DO (∆DXA = ∆3DO (slope) + intercept)

Bolded mean differences signify a p-value < 0.05

a

Supplemental Table 2. Adjustments to change in 3DO and change in DXA relationship

| Outcome | Variables | Beta | R^2^ | RMSE |
| --- | --- | --- | --- | --- |
| Female ∆Total FM | Intercept | 0.109 | 0.90 | 1.67 |
|  | ∆3DO Fat | 0.420 |  |  |
|  | ∆Weight | 0.418 |  |  |
| Female ∆Total FFM | Intercept | -0.023 | 0.80 | 1.35 |
|  | ∆3DO LM | 0.035 |  |  |
|  | ∆Weight | 0.317 |  |  |
| Female ∆%Fat | Intercept | -0.046 | 0.51 | 1.77 |
|  | ∆3DO %Fat | 0.300 |  |  |
|  | ∆BMI | 0.513 |  |  |
| Female ∆ALM | Intercept | 0.657 | 0.79 | 0.31 |
|  | ∆3DO ALM | 0.264 |  |  |
|  | ∆Weight | 0.038 |  |  |
|  | BL Weight | -0.008 |  |  |
| Female ∆VAT | Intercept | 0.016 | 0.31 | 0.09 |
|  | ∆3DO VAT | 0.736 |  |  |
| Male ∆Total FM | Intercept | -20.58 | 0.80 | 2.01 |
|  | ∆3DO Fat | 1.243 |  |  |
|  | BL Height | 0.112 |  |  |
|  | Ethnicity (W) | Ref |  |  |
|  | Ethnicity (B) | -0.184 |  |  |
|  | Ethnicity (H) | 3.357 |  |  |
|  | Ethnicity (A) | 0.991 |  |  |
| Male ∆Total FFM | Intercept | -0.037 | 0.76 | 1.69 |
|  | ∆3DO LM | 0.580 |  |  |
|  | ∆Weight | 0.063 |  |  |
| Male ∆%Fat | Intercept | -20.15 | 0.42 | 2.01 |
|  | ∆3DO %Fat | 0.645 |  |  |
|  | ∆Weight | 0.077 |  |  |
|  | Ethnicity (W) | Ref |  |  |
|  | Ethnicity (B) | -0.122 |  |  |
|  | Ethnicity (H) | 3.157 |  |  |
|  | Ethnicity (A) | 1.570 |  |  |
|  | BL Height | 0.104 |  |  |
| Male ∆ALM | Intercept | -0.019 | 0.56 | 0.48 |
|  | ∆3DO ALM | 0.402 |  |  |
|  | ∆Weight | 0.025 |  |  |
| Female ∆VAT | Intercept | 0.141 | 0.28 | 0.11 |
|  | ∆3DO VAT | 0.560 |  |  |
|  | BL BMI | -0.006 |  |  |

Abbreviations: ALM (appendicular lean mass), BL (Baseline), ∆ (Difference between follow-up and baseline), FM (fat mass), LM (lean mass), %Fat (percent fat), W (White), B (Black), A (Asian), H (Hispanic)

The outcome variables are from DXA, and the predictor variables are from 3DO and demographics.

Supplemental Table 3. Agreement between 3DO and DXA for those exceeding and not exceeding significant change with unadjusted estimates.

|  | **Female** | **Male** | **Overall** |
| --- | --- | --- | --- |
|  | **(N=45)** | **(N=88)** | **(N=133)** |
| **Total Fat Mass** |  |  |  |
| 3DO^1^ | 1 (2.2%) | 6 (6.8%) | 7 (5.3%) |
| DXA^2^ | 15 (33.3%) | 19 (21.6%) | 34 (25.6%) |
| Both Sig/Not Sig^3^ | 29 (64.5%) | 63 (71.6%) | 92 (69.1%) |
| Kappa Score | 0.39 | 0.42 | 0.41 |
| **Total Fat-free Mass** |  |  |  |
| 3DO^1^ | 2 (4.4%) | 5 (5.7%) | 7 (5.3%) |
| DXA^2^ | 9 (20.0%) | 13 (14.8%) | 22 (16.5%) |
| Both Sig/Not Sig^3^ | 34 (75.6%) | 70 (79.5%) | 104 (78.2%) |
| Kappa Score | 0.43 | 0.44 | 0.44 |
| **Percent Fat** |  |  |  |
| 3DO^1^ | 1 (2.2%) | 4 (4.5%) | 5 (3.8%) |
| DXA^2^ | 18 (40.0%) | 40 (45.5%) | 58 (43.6%) |
| Both Sig/Not Sig^3^ | 26 (57.8%) | 44 (50.0%) | 70 (52.6%) |
| Kappa Score | 0.37 | 0.33 | 0.34 |
| **ALM** |  |  |  |
| 3DO^1^ | 3 (6.7%) | 11 (12.5%) | 14 (10.5%) |
| DXA^2^ | 4 (8.9%) | 12 (13.6%) | 16 (12.0%) |
| Both Sig/Not Sig^3^ | 38 (84.4%) | 65 (73.9%) | 103 (77.5%) |
| Kappa Score | 0.46 | 0.42 | 0.43 |
| **VAT** |  |  |  |
| 3DO^1^ | 14 (31.1%) | 18 (20.5%) | 32 (24.1%) |
| DXA^2^ | 6 (13.3%) | 8 (9.1%) | 14 (10.5%) |
| Both Sig/Not Sig^3^ | 25 (55.6%) | 62 (70.4%) | 87 (65.4%) |
| Kappa Score | 0.36 | 0.41 | 0.39 |

Abbreviations: ALM (appendicular lean mass), LSC (least significant change), Sig (significant), VAT (visceral adipose tissue)

^1^ Participants that surpassed the LSC for only 3DO.

^2^ Participants that surpassed the LSC for only DXA.

^3^ Participants that surpassed both the LSC for 3DO and DXA or did not detect a significant change on either 3DO or DXA.

Supplemental Table 4. (Females) Comparison of different modeling techniques for 3DO vs DXA body composition change.

| **Models** | **Outcome** | **Variables** | **Beta** | **R^2^** | **RMSE** |
| --- | --- | --- | --- | --- | --- |
| 1, 2, 3 | ∆Total FM | Intercept | 0.103 | 0.94 | 1.29 |
|  |  | ∆PC2 | 2.524 |  |  |
|  |  | ∆PC3 | -1.417 |  |  |
|  |  | ∆PC4 | -1.105 |  |  |
|  |  | ∆PC10 | 2.345 |  |  |
|  |  | ∆PC12 | 2.987 |  |  |
|  |  | ∆PC13 | -2.996 |  |  |
|  | ∆Total FFM | ∆Weight - ∆FM |  | 0.82 | 1.26 |
|  | ∆%Fat | ((∆FM*BL Weight) + (BL FM*∆Weight) / (BL Weight*FU Weight)) *100 |  | 0.64 | 1.49 |
| 4 | ∆Total FM | Intercept | -0.867 | 0.9 | 1.53 |
|  |  | BL PC2 | -2.243 |  |  |
|  |  | BL PC3 | 1.669 |  |  |
|  |  | BL PC12 | -1.194 |  |  |
|  |  | BL PC13 | 2.5873 |  |  |
|  |  | FU PC2 | 2.224 |  |  |
|  |  | FU PC3 | -1.792 |  |  |
|  |  | FU PC9 | -1.172 |  |  |
|  |  | FU PC13 | -4.351 |  |  |
|  | ∆Total FFM | ∆Weight - ∆FM |  | 0.77 | 1.53 |
|  | ∆%Fat | ((∆FM*BL Weight) + (BL FM*∆Weight) / (BL Weight*FU Weight)) *100 |  | 0.52 | 1.7 |
| 5 | ∆Total FM | Intercept | -0.315 | 0.9 | 1.66 |
|  |  | ∆PC3 | 0.399 |  |  |
|  |  | ∆Weight | 0.612 |  |  |
|  | ∆Total FFM | ∆Weight - ∆FM |  | 0.71 | 1.6 |
|  | ∆%Fat | ((∆FM*BL Weight) + (BL FM*∆Weight) / (BL Weight* FU Weight)) *100 |  | 0.39 | 1.73 |
| 5 | ∆VAT | Intercept | 0.014 | 0.43 | 0.08 |
|  |  | ∆PC11 | -0.076 |  |  |
|  |  | ∆Weight | 0.008 |  |  |

Abbreviations: BL (Baseline), FU (follow-up), ∆ (Difference between follow-up and baseline), FM (fat mass), FFM (fat-free mass), %Fat (percent fat)

Models were built with step forward linear regression with 5-fold cross validation. The outcome variables are from DXA, and the predictor variables are from 3DO.

Model 1: ∆ PCs; Model 2: model 1 + baseline total fat mass; Model 3: model 1 + baseline PCs; Model 4: baseline and follow-up PCs; Model 5: model 1+ change in weight

Best model for ∆VAT presented.

Supplemental Table 5. (Males) Comparison of different modeling techniques for 3DO vs DXA body composition change.

| Model | Outcome | Variables | Beta | R^2^ | RMSE |
| --- | --- | --- | --- | --- | --- |
| 1, 2, 3 | ∆Total FM | Intercept | -0.313 | 0.84 | 1.79 |
|  |  | ∆PC1 | -0.279 |  |  |
|  |  | ∆PC2 | 2.097 |  |  |
|  |  | ∆PC3 | -1.338 |  |  |
|  |  | ∆PC5 | -1.1 |  |  |
|  |  | ∆PC12 | 3.11 |  |  |
|  | ∆Total FFM | ∆Weight - ∆FM |  | 0.75 | 1.76 |
|  | ∆%Fat | ((∆FM*BL Weight) + (BL FM*∆Weight) / (BL Weight*FU Weight)) *100 |  | 0.45 | 2 |
| 4 | ∆Total FM | Intercept | -2.248 | 0.72 | 2.37 |
|  |  | BL PC2 | -1.372 |  |  |
|  |  | BL PC6 | 0.695 |  |  |
|  |  | BL PC10 | 2.251 |  |  |
|  |  | BL PC11 | 2.184 |  |  |
|  |  | FU PC2 | 1.196 |  |  |
|  |  | FU PC5 | -0.764 |  |  |
|  |  | FU PC13 | -2.836 |  |  |
|  | ∆Total FFM | ∆Weight - ∆FM |  | 0.7 | 2.36 |
|  | ∆%Fat | ((∆FM*BL Weight) + (BL FM*∆Weight) / (BL Weight*FU Weight)) *100 |  | 0.27 | 2.24 |
| 5 | ∆Total FM | Intercept | 0.02 | 0.87 | 1.66 |
|  |  | ∆Weight | 0.595 |  |  |
|  | ∆Total FFM | ∆Weight - ∆FM |  | 0.75 | 1.52 |
|  | ∆%Fat | ((∆FM*BL Weight) + (BL FM*∆Weight) / (BL Weight*FU Weight)) *100 |  | 0.59 | 1.48 |
| 5 | ∆VAT | Intercept | -0.046 | 0.36 | 0.10 |
|  |  | ∆PC2 | 0.022 |  |  |
|  |  | ∆PC9 | -0.076 |  |  |
|  |  | ∆PC12 | 0.106 |  |  |
|  |  | BL PC5 | -0.028 |  |  |
|  |  | BL PC14 | -0.077 |  |  |

Abbreviations: BL (Baseline), FU (follow-up), ∆ (Difference between follow-up and baseline), FM (fat mass), FFM (fat-free mass), %Fat (percent fat), VAT (visceral adipose tissue)

Models were built with step forward linear regression with 5-fold cross validation. The outcome variables are from DXA, and predictor variables are from 3DO and demographics.

Model 1: ∆ PCs; Model 2: model 1 + baseline total fat mass; Model 3: model 1 + baseline PCs; Model 4: baseline and follow-up PCs; Model 5: model 1+ change in weight

Best model for ∆VAT presented.

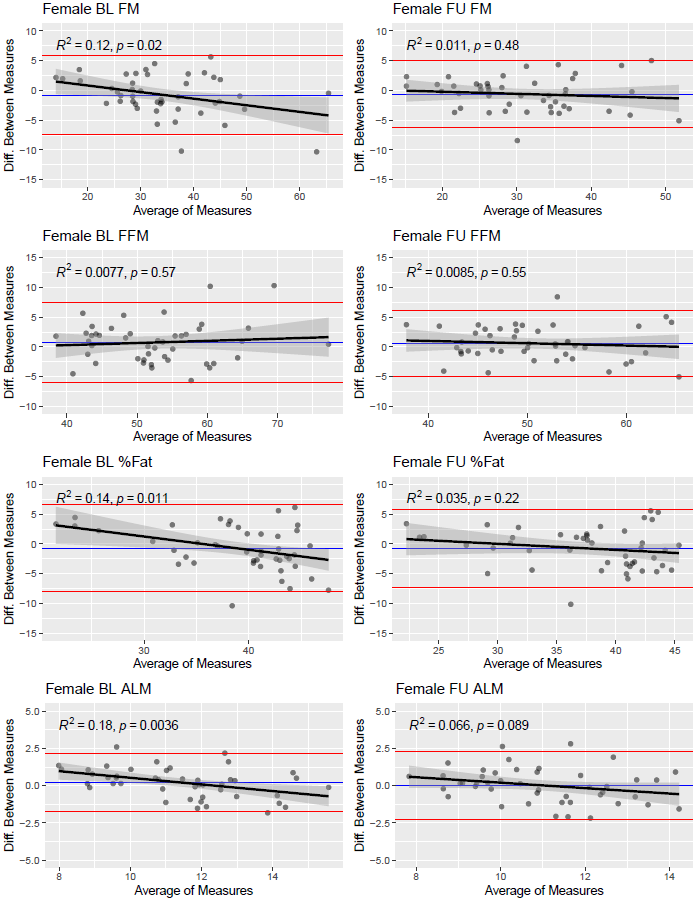

Supplemental Figure 2. Female baseline and follow-up Bland-Altman plots comparing DXA and 3DO body composition. Mean difference (blue line), ± 1.96 x standard deviation (red lines), and best fit line (black line) are shown on each plot. All differences on y-axis are 3DO – DXA. Abbreviations: ALM (appendicular lean mass), BL (baseline), FM (fat mass), FFM (fat-free mass), FU (follow-up).

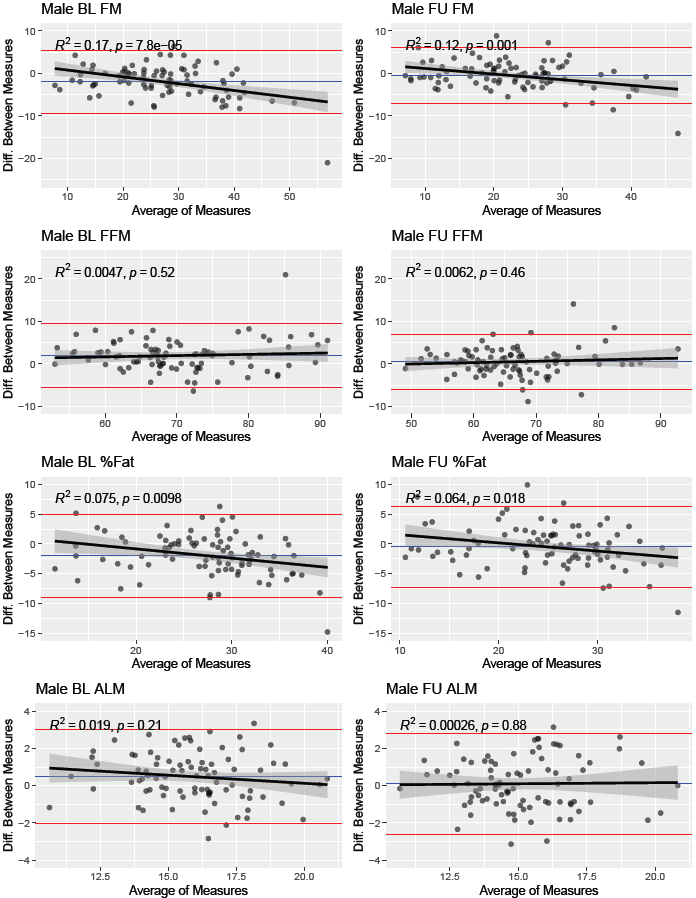

Supplemental Figure 3. Male baseline and follow-up Bland-Altman plots comparing DXA and 3DO. Mean difference (blue line), ± 1.96 x standard deviation (red lines), and best fit line (black line) are shown on each plot. All differences on y-axis are 3DO – DXA. Abbreviations: ALM (appendicular lean mass), BL (baseline), FM (fat mass), FFM (fat-free mass), FU (follow-up).
